# Estimating the infection fatality risk of COVID-19 in New York City during the spring 2020 pandemic wave

**DOI:** 10.1101/2020.06.27.20141689

**Authors:** Wan Yang, Sasikiran Kandula, Mary Huynh, Sharon K. Greene, Gretchen Van Wye, Wenhui Li, Hiu Tai Chan, Emily McGibbon, Alice Yeung, Don Olson, Anne Fine, Jeffrey Shaman

## Abstract

**Background:** As the COVID-19 pandemic continues to unfold, the infection fatality risk (IFR, i.e. risk of death among all infections including asymptomatic and mild infections) is crucial for gauging the burden of death due to COVID-19 in the coming months or years. Here we estimate the IFR of COVID-19 in New York City (NYC), the first epidemic center in the United States where the IFR remains unclear.

**Methods:** We developed a meta-population network model-inference system to estimate underlying SARS-CoV-2 infection rates in NYC during the 2020 spring pandemic wave using case, mortality, and mobility data. Based on these estimates, we further estimated the IFR for all ages overall and for 5 age groups (i.e. <25, 25-44, 45-64, 65-74, and 75+ years) separately, during March 1 – June 6, 2020 (i.e., before NYC began its phased reopening).

**Findings:** During March 1 – June 6, 2020, 205,639 laboratory-confirmed COVID-19 cases were diagnosed and 21,447 confirmed and probable COVID-19 deaths occurred among NYC residents. We estimated an overall IFR of 1.39% (95% Credible Interval: 1.04-1.77%) in NYC. Estimated IFR for the two oldest age groups (65-74 and 75+ years) was much higher than the younger age groups and about double estimates reported for elsewhere. In particular, weekly IFR was estimated as high as 6.7% for 65-74 year-olds and 19.1% for 75+ year-olds.

**Interpretation:** These results are based on more complete ascertainment of COVID-19-associated deaths in NYC and thus likely more accurately reflect the true, higher burden of death due to COVID-19 than previously reported elsewhere.

## Introduction

The novel coronavirus SARS-CoV-2 emerged in late 2019 in China and subsequently spread to 200+ other countries. As of Aug 8, 2020, there were over 19 million reported COVID-19 cases and over 700 thousand deaths worldwide.^1^ As the pandemic continues to unfold and populations in many places worldwide largely remain susceptible, understanding the severity, in particular, the infection fatality risk (IFR), is crucial for gauging the full impact of COVID-19 in the coming months or years. However, estimating the IFR of COVID-19 is challenging due to the large number of undocumented infections, fluctuating infection detection rates, and inconsistent reporting of fatalities. Further, the IFR of COVID-19 could vary by location, given differences in demographics, healthcare systems, and social construct (e.g., intergenerational households are the norm in some societies whereas older adults commonly reside and congregate in long-term care and adult care facilities in others). Most IFR estimates thus far have come from data recorded in China, the Diamond Princess cruise ship, and France.^2-5^ To date, the IFR in the United States—the country currently reporting the largest number of cases—remains unclear.

New York City (NYC) reported its first COVID-19 case on March 1, 2020, in a traveler, and quickly became the epicenter in the United States. Intense community transmission occurred during the following three months before a series of public health interventions brought the pandemic under control. By June 6, 2020, prior to the city’s reopening, there were 205,639 diagnosed cases and 21,447 deaths reported in NYC (Table 1). During the pandemic, the NYC Department of Health and Mental Hygiene (DOHMH) and the Mailman School of Public Health at Columbia University have been collaborating in generating real-time model projections in support of the city’s pandemic response. Our latest model-inference system uses a meta-population network model to simulate SARS-CoV-2 transmission in the city’s 42 United Hospital Fund neighborhoods.^6^ The model is run in conjunction with the Ensemble Adjustment Kalman Filter (EAKF)^7^ and fit simultaneously to case and mortality data for each of the 42 neighborhoods while accounting for under-detection, delay from infection to case reporting and death, and changing interventions (e.g., social distancing). In this study, we apply this network model-inference system to estimate the IFR for 5 age groups (i.e. <25, 25-44, 45-64, 65-74, and 75+ years) and all ages overall, from March 1 to June 6, 2020. In the process, we also estimate infection detection rates—i.e. the fraction of infections documented as confirmed cases—and the cumulative infection rate by June 6, 2020.

**Table 1.** Summary estimates. Cases and deaths were reported during March 1 – June 6, 2020. Cumulative infection rates, median (95% CrI), show percentages of population, for each age group or all ages overall, estimated to have been infected by June 6, 2020. IFR, median (95% CrI), was estimated here, and averaged over March 22 – June 6, 2020; we excluded estimates during March 1-21, 2020, because estimates were less accurate for these earliest weeks when zero or few deaths were reported.

| Age | Confirmed | Confirmed and | Estimated cumulative |  |
| --- | --- | --- | --- | --- |
|  | Cases | Probable Deaths | infection rate (%) | Estimated IFR (%) |
| <25 | 16332 | 45 | 8.6 (5.7, 17.5) | 0.0097 (0.0041, 0.0154) |
| 25-44 | 64753 | 734 | 22.6 (16.6, 31.2) | 0.12 (0.073, 0.15) |
| 45-64 | 74798 | 4732 | 22.7 (18, 29.2) | 0.94 (0.73, 1.19) |
| 65-74 | 25460 | 5181 | 15.0 (11.4, 21.6) | 4.87 (3.37, 6.89) |
| 75+ | 24296 | 10755 | 12.8 (9.9, 18.6) | 14.17 (10.18, 18.14) |
| all | 205639 | 21447 | 17.2 (12.9, 25.1) | 1.39 (1.04, 1.77) |

## Methods

### Data

Laboratory-confirmed COVID-19 cases reported to the NYC DOHMH were aggregated by week of diagnosis and age group (<1, 1-4, 5-14, 15-24, 25-44, 45-64, 65-74, and 75+ years) for each of the 42 United Hospital Fund neighborhoods^6^ in NYC, according to the patient’s residential address at time of report. The mortality data, from deaths registered and analyzed by the NYC DOHMH, combined confirmed and probable COVID-19-associated deaths. Confirmed COVID-19-associated deaths were defined as those occurring in persons with laboratory-confirmed SARS-CoV-2 infection; and probable COVID-19 deaths were defined as those with COVID-19, SARS-CoV-2, or a similar term listed on the death certificate as an immediate, underlying, or contributing cause of death but did not have laboratory-confirmation of COVID-19.^8^ Due to privacy concerns, mortality data were aggregated to 5 coarser age groups (<18, 18-44, 45-64, 65-74, and 75+ years) for each neighborhood by week of death. To match with the age grouping for case data, we used the citywide fraction of deaths occurring in each of the five finer age groups (i.e. <1, 1-4, 5-14, 15-24, 25-44) to apportion deaths in the <18 and 18-44 year age categories. For this study, case and mortality data were both retrieved on August 7, 2020.

The mobility data, used to model changes in COVID-19 transmission rate due to public health interventions implemented during the pandemic (e.g., social distancing), came from SafeGraph^9,10^ and contained counts of visitors to locations in each zip code based on mobile device locations. The released data were anonymized and aggregated in weekly intervals. We spatially aggregated these data to the neighborhood level.

This study was classified as public health surveillance and exempt from ethical review and informed consent by the Institutional Review Boards of both Columbia University and NYC DOHMH.

### Meta-population network transmission model

The meta-population network model simulated intra- and inter neighborhood transmission of COVID-19 and assumed susceptible-exposed-infectious-removed (SEIR) dynamics, per the following equations:

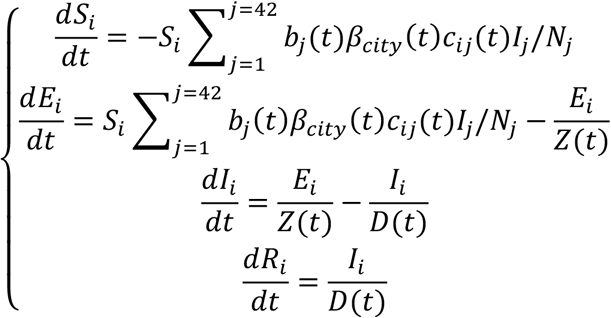

where *S_i_, E_i_, I_i_, R_i_*, and *N_i_* are the numbers of susceptible, exposed (but not yet infectious), infectious, and removed (either recovered or deceased) individuals and the total population, respectively, from a given age group in neighborhood *i*. Note that due to model complexity and a lack of information for parameterizing interactions among age groups, we modeled each age group separately (i.e., combining all sources of infection to each age group; see further detail on parameter estimation below); as such, Eqn 1 describes the spatial transmission across neighborhoods with no interactions among age groups. *t* is time; we make this time dependence explicit for the parameters to indicate that they were estimated for each week (see below) and could vary over time due to disease seasonality and/or public health interventions; state variables (*S_i_*, *E_i_, I_i_* and *R_i_*) are inherently time-varying. *β_city_*(*t*) is the citywide transmission rate, which incorporated seasonal variation as observed for OC43, a beta-coronavirus in humans from the same genus as SARS-CoV-2 (see details in the Appendix, pages 1-3). To allow differential transmission in each neighborhood, we included a multiplicative factor, *b_i_*, to scale neighborhood local transmission rates. *Z* and *D* are the latency and infectious periods, respectively (Appendix, Table S1).

The matrix [*c_ij_*(*t*)] represents changes in contact rates over time and connectivity among neighborhoods and was computed based on mobility data. Briefly, changes in contact rates (either intra or inter neighborhoods) for week-t were computed as a ratio of the number of visitors during week-t to that during the week of March 1, 2020 (the first week of the pandemic in NYC when there were no interventions in place), and further scaled by a multiplicative factor *m_1_; m_1_* was estimated along with other parameters. To compute the connectivity among the neighborhoods, we first divided the inter-neighborhood mobility by the local mobility (this gave a relative measure of connectivity; e.g., if two neighborhoods are highly connected with lots of individuals traveling between them, inter-neighborhood mobility would be closer to 1 and much lower than 1 otherwise); we then scaled these relative rates by a multiplicative factor *m_2_*, which was also estimated along with other parameters.

### Observation model

To account for delays in diagnosis and detection, we included a lag of time-from-infectious-to-detection (i.e., an infection being diagnosed as a case), drawn from a gamma distribution with a mean of *T_m_* and standard deviation (SD) of *T_sd_* days. To account for under-detection, we included an infection detection rate (*r*), i.e. the fraction of infections (including subclinical or asymptomatic infections) reported as cases. To compute the model-simulated number of new cases per week, we multiplied the model-simulated number of infections per day (including those from the previous weeks) by the infection detection rate, and further distributed these simulated cases in time per the distribution of time-from-infectious-to-detection. We then aggregated the daily lagged, simulated cases to weekly totals for model inference (see below). Similarly, to compute the model-simulated deaths per week and account for delays in time to death, we multiplied the simulated-infections by the IFR and then distributed these simulated deaths in time per the distribution of time-from-infectious-to-death, and aggregated these daily numbers to weekly totals. For each week, the infection detection rate (*r*), the mean (*T_m_*) and standard deviation (*T_sd_*) of time-from-infectious-to-detection, and the IFR were estimated based on weekly case and mortality data. The distribution of time-from-diagnosis-to-death was based on observations of *n*=15,686 COVID-19 confirmed deaths in NYC (gamma distribution with mean = 9.36 days and SD = 9.76 days; Appendix, Table S1).

### Parameter estimation

To estimate model parameters (*β_city_, Z, D, m_1_, m_2_, T_m_, T_sd_, r, IFR* and *b_i_*, for *i*=1,…,42) and state variables (*S_i_*, *E_i_*, and *I_i_*, for *i*=1,…,42) for each week, we ran the meta-population network-model stochastically with a daily time step in conjunction with the EAKF and fit to weekly case and mortality data from the week starting March 1 to the week ending June 6, 2020. The EAKF uses an ensemble of model realizations (*n*=500 here), each with initial parameters and variables randomly drawn from a prior range (see Appendix, Table S1). After model initialization, the model ensemble was integrated forward in time for a week to compute the model-simulated number of cases and deaths for that week; these prior estimates were then combined with the observed cases and deaths for the same week to compute the posterior per Bayes’ theorem.^7^ Of note, the EAKF also models the observational errors (e.g., due to imperfect sensitivity and specificity of RT-PCR test for case diagnosis) over time by specifying an error structure and using this information when computing the posterior.^7^ The posterior distribution of each model parameter/variable was updated for that week at the same time (note that stationary parameters would remain so after converging to their solutions).^7^ This parameter estimation process was done separately for each of the eight age groups (i.e. <1, 1-4, …, and 75+). To include transmission from other age groups, we used measured intra and inter-group contacts from the POLYMOD study^11^ to compute the total number of contacts made to each age group and adjusted the *prior* range of the transmission rate (*β_city_*) during the first week of the pandemic for each age group accordingly. The posterior estimate was computed based on case and mortality data for each group, which included all sources of infection. Thus, the estimated transmission rate for each age group nevertheless included all sources of transmission.

To account for stochasticity in model initiation, we ran the parameter estimation process independently 10 times. Results for each age group were combined from these 10 runs (each with 500 realizations). To combine estimates of infection detection rate and IFR for <25 year-olds or all ages overall, we weighted the age-group specific estimates [median and credible interval (CrI)] by the fraction of estimated infections from each related group.

## Results

The model-inference system was able to recreate the case and mortality time series for each age group and all ages overall (Fig. 1). For most age groups, confirmed cases peaked during the week of March 29 and the mortality rate peaked about one week later than the case rate, due to the time-lag from severe infection to death (Fig. 1).

**Figure 1.**
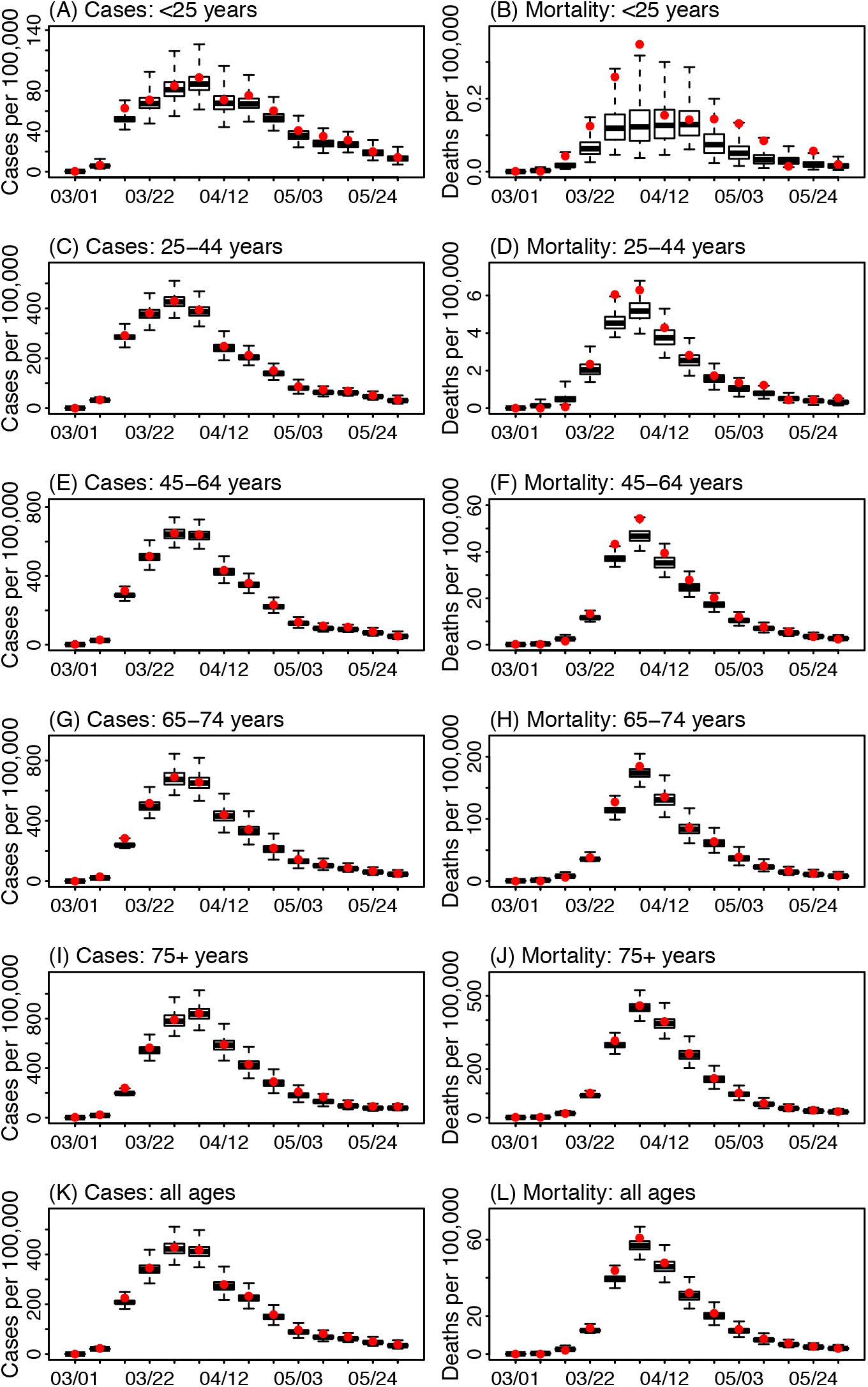
Model fit. Left panel: model estimates of confirmed COVID-19 cases per 100,000 population for (A) <25 year-olds, (C) 25-44 year-olds, (E) 45-64 year-olds, (G) 65-74 year-olds, (I) 75+ year-olds, and (K) all ages overall. Right panel: model estimates of COVID-19 associated deaths per 100,000 population for (B) <25 year-olds, (D) 25-44 year-olds, (F) 45-64 year-olds, (H) 65-74 year-olds, (J) 75+ year-olds, and (L) all ages overall. Boxes and whiskers represent median, 50% CrI, and 95% Crl. Red dots represent observed confirmed case rates (left panel) and observed mortality rates (right panel). x-axis shows the first day of each week (mm/dd) from the week of March 1 to the week of May 31, 2020.

There were, however, substantial under-detection of infections, variations by age group, and fluctuations of infection detection rates over time, in part due to changing testing criteria.^12,13^ The estimated infection detection rate for all ages overall started at a low level of 2.2% [median; 95% CrI: 0.3–4.5%; same below] in the week of March 1; it increased to 17.4% (95% CrI: 11.3–26.1%) during the week of March 15. However, due to material shortages in testing equipment and personal protective equipment, testing was restricted to severely ill patients in early April^12^ before it became more widely available in May.^13^ Consistently, the estimated infection detection rate dropped to ~13% in early-mid April, then gradually increased to ~19% in early May and stayed at similar levels through the week of May 31, 2020 (Fig 2F).

**Figure 2.**
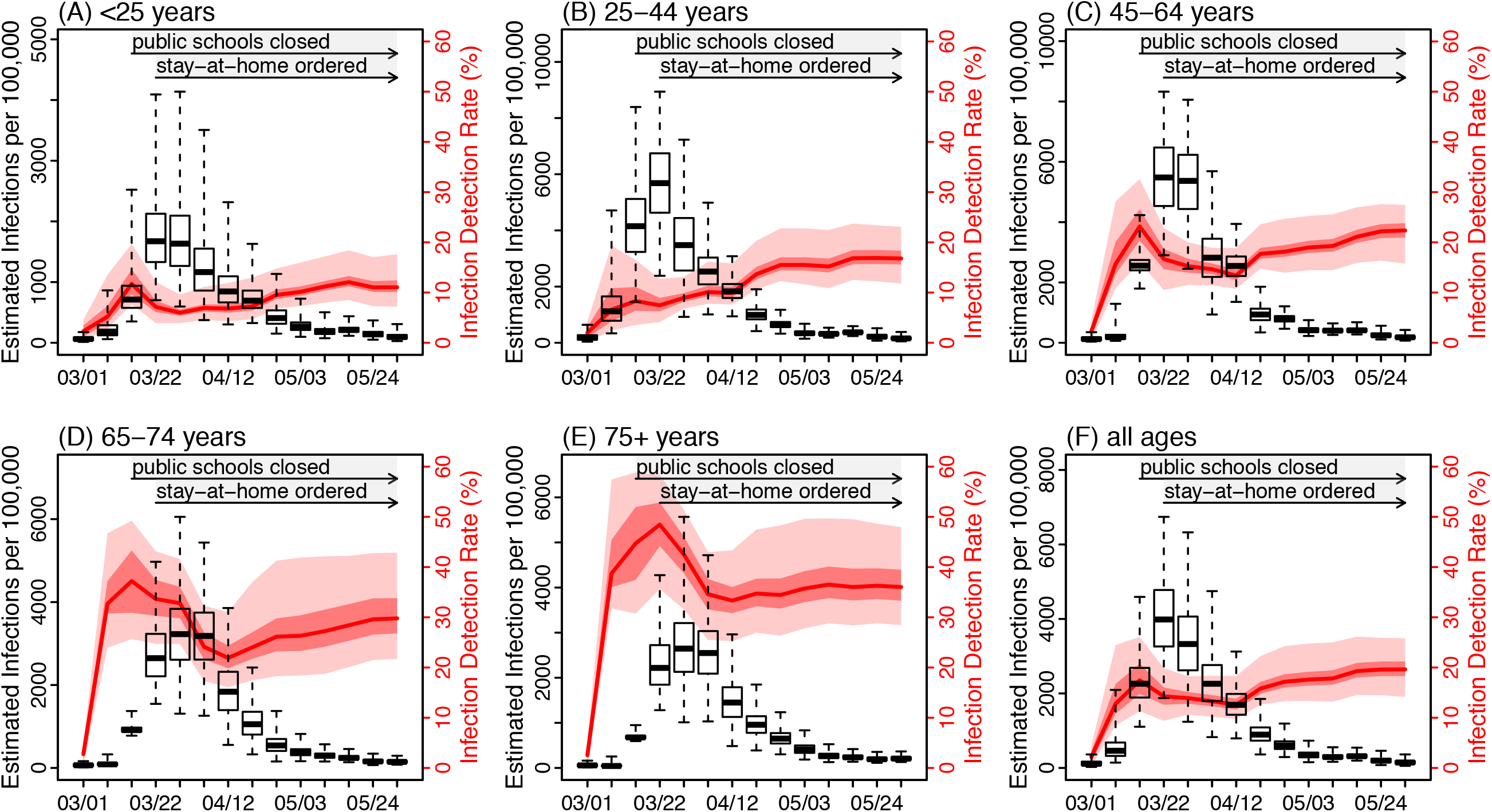
Estimated infection rates and infection detection rates over time for (A) <25 year-olds, (B) 25-44 year-olds, (C) 45-64 year-olds, (D) 65-74 year-olds, (E) 75+ year-olds, and (F) all ages overall. Boxes and whiskers show estimated infection rates (median, 50% CrI, and 95% CrI; left y-axis). Red lines and surrounding areas show the estimated infection detection rates (median, 50% CrI, and 95% CrI; right y-axis). x-axis shows the first day of each week (mm/dd) from the week of March 1 to the week of May 31, 2020. Horizontal arrows and grey shaded areas indicate the timing of two major public health intervention measures, i.e., school closures starting the week of March 15, 2020 and stay-at-home mandate starting the week of March 22, 2020.

The estimated infection detection rate was highest for the two oldest age groups and substantially lower for younger age groups (Fig 2 D and E vs. Fig 2 A-C). During the week of May 31, 2020 prior to the city began its phased reopening, we estimated that 29.8% (95% CrI: 21.7–42.3%) of infections among 65-74 year-olds and 36.0% (95% CrI: 28.4–47.9%) among 75+ year-olds were detected; in comparison, only 11.0% (95% CrI: 7.2–17.6%) of infections among <25 year-olds and 16.8% (95% CrI: 11.8–23.1%) among 25-44 year-olds were detected.

After accounting for the infection detection rate, the epidemic peak for new infections occurred 1-2 weeks sooner during the week of March 22, 2020 for <65 year-olds and all ages combined (Fig 2 A-C and F). This was coincident with the timing of public health interventions in NYC – public schools in NYC were closed on March 16, 2020 and a citywide stay-at-home order was imposed starting the week of March 22, 2020.^14^ Tallied over the entire study period, the estimated overall cumulative infection rate was 17.2% (95% CrI: 12.9–25.1%) by June 6, 2020. However, the estimated cumulative infection rates varied substantially across age groups and neighborhoods in NYC (Fig 3). Specifically, 25-44 and 45-64 year-olds had the highest cumulative infection rates, at 22.6% (95% CrI: 16.6–31.2%) and 22.7% (95% CrI: 18.0–29.2%), respectively; 65-74 and 75+ year-olds had the second highest cumulative infection rates, at 15.0% (95% CrI: 11.4–21.6%) and 12.8% (95% CrI: 9.9–18.6%); and <25 year-olds had the lowest cumulative infection rate (8.6%; 95% CrI: 5.7–17.5%). Spatially, among the five boroughs in NYC (i.e. Manhattan, Bronx, Brooklyn, Queens, and Staten Island), estimated cumulative infection rates were highest in neighborhoods in the Bronx and lowest in neighborhoods in Manhattan (Fig 3).

**Figure 3.**
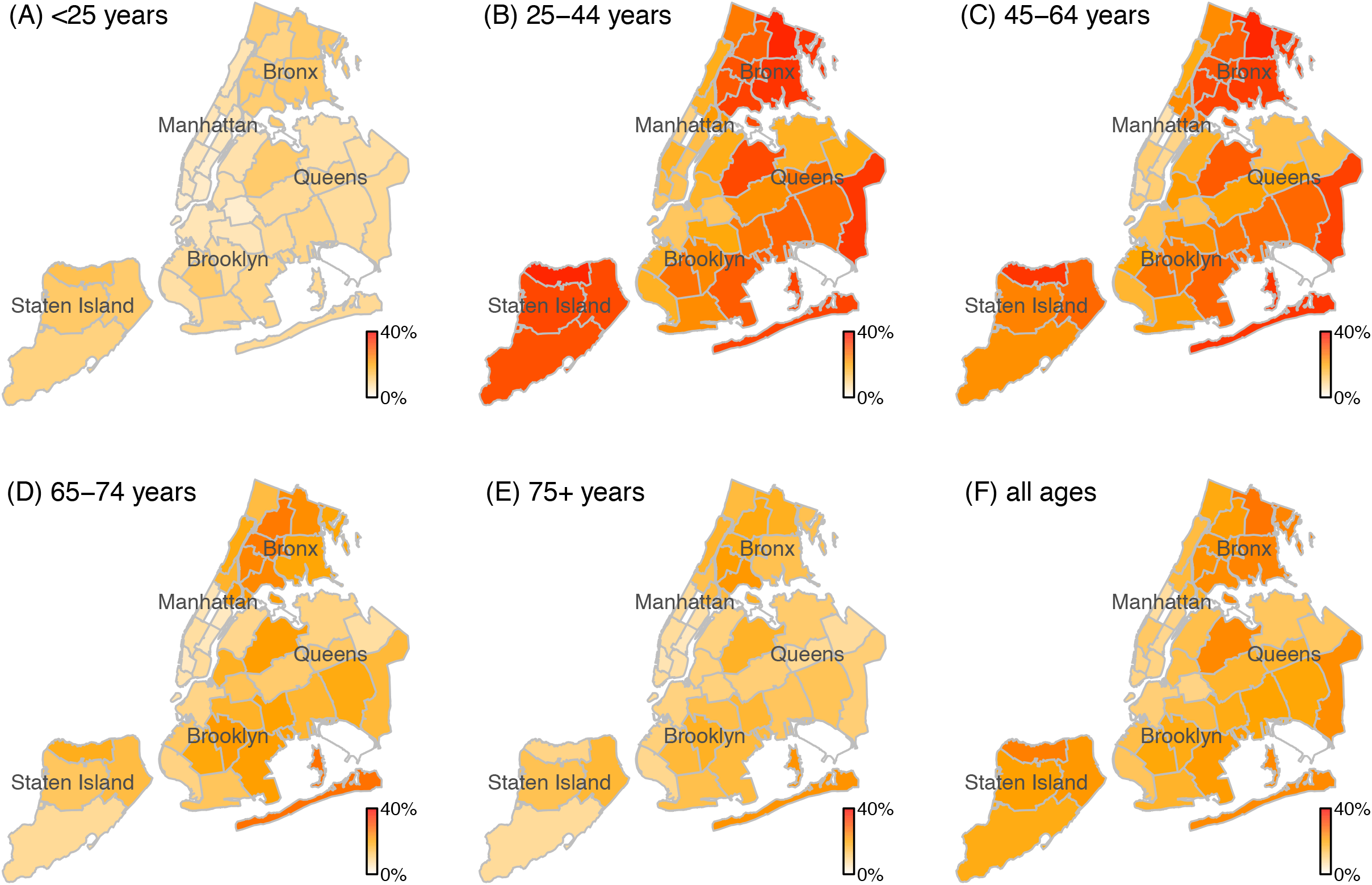
Estimated cumulative infection rates across neighborhoods in NYC among (A) <25 year-olds, (B) 25-44 year-olds, (C) 45-64 year-olds, (D) 65-74 year-olds, (E) 75+ year-olds, and (F) all ages overall. NYC has five boroughs (i.e. Manhattan, Bronx, Brooklyn, Queens, and Staten Island; labeled on the maps) and 42 neighborhoods (delineated by the light grey lines). The heat maps show the estimated median cumulative infection rates (March 1 – June 6, 2020) for each age group and neighborhood.

There are large uncertainties in our model estimates of cumulative infection rates. To evaluate the accuracy, we compared our model estimates with three datasets of seroprevalence of antibodies to SARS-CoV-2 measured during three phases of the pandemic in NYC (i.e. early-phase in March,^15^ mid-phase in April,^16^ and end of the pandemic;^17^ see details on available serology data and matching by timing of measurement, age group, and location in the Appendix, page 1 and Table S2). As shown in Appendix, Fig S1, albeit with large uncertainties, our estimated cumulative infection rates were in line with corresponding measures from antibody tests, for all three phases of the pandemic wave. Consistent with serology data, our model-inference system estimated higher infection rates among adults 25-64 years than other age groups. In addition, the spatial variation estimated by our model-inference system was also in line with reported measures (i.e., highest in the Bronx and lowest in Manhattan). This consistency with independent serology data provides some independent validation of our model estimates.

During March 1 – June 6, 2020, a total of 21,447 COVID-19-associated deaths (16,924 confirmed and 4,523 probable) occurred and 205,639 COVID-19 cases were diagnosed in NYC. The crude confirmed case fatality risk was thus 8.23%. After accounting for changing infection detection rates and excluding the first three weeks (i.e., March 1-21, 2020) with zero or few reported deaths for which model estimates were less accurate, we estimate that the overall IFR, including both confirmed and probable deaths, was 1.39% (95% CrI: 1.04–1.77%) during March 22 – June 6, 2020.

Examining estimates by age group, estimated IFR was lowest in young age groups. The average IFR was 0.0097% (95% CrI: 0.0041–0.015%) for <25 year-olds, increased by ~10 fold to 0.12% (95% CrI: 0.073–0.15%) for 25-44 year-olds, and by another 8 fold to 0.94% (95% CrI:0.73–1.19%) for 45-64 year-olds (Fig. 4 A-C). These estimates were similar to IFRs reported for China for corresponding age groups.^3^ However, the estimated IFR for the two oldest age groups was much higher than the younger age groups and about twice as high as rates reported for these age groups in China.^3,4^ The average IFR was 4.87% (95% CrI: 3.37–6.89%) for 65-74 year-olds and 14.17% (95% CrI: 10.18–18.14%) for 75+ year-olds. In addition, the estimated IFR fluctuated substantially over time for these two elderly groups. For 65-74 year-olds, estimated IFR was 6.72% (95% CrI: 5.52–8.01%) during the week of April 5, 2020 but decreased to 4.20% (95% CrI: 2.22–7.01%) during the week of May 31, 2020 (Fig 4D). For 75+ year-olds, IFR was estimated to be 19.11% (95% CrI: 14.70–21.92%) during the week of April 5, 2020 but decreased to 10.38% (95% CrI: 6.17–14.96%) during the week of May 31, 2020 (Fig 4F).

**Figure 4.**
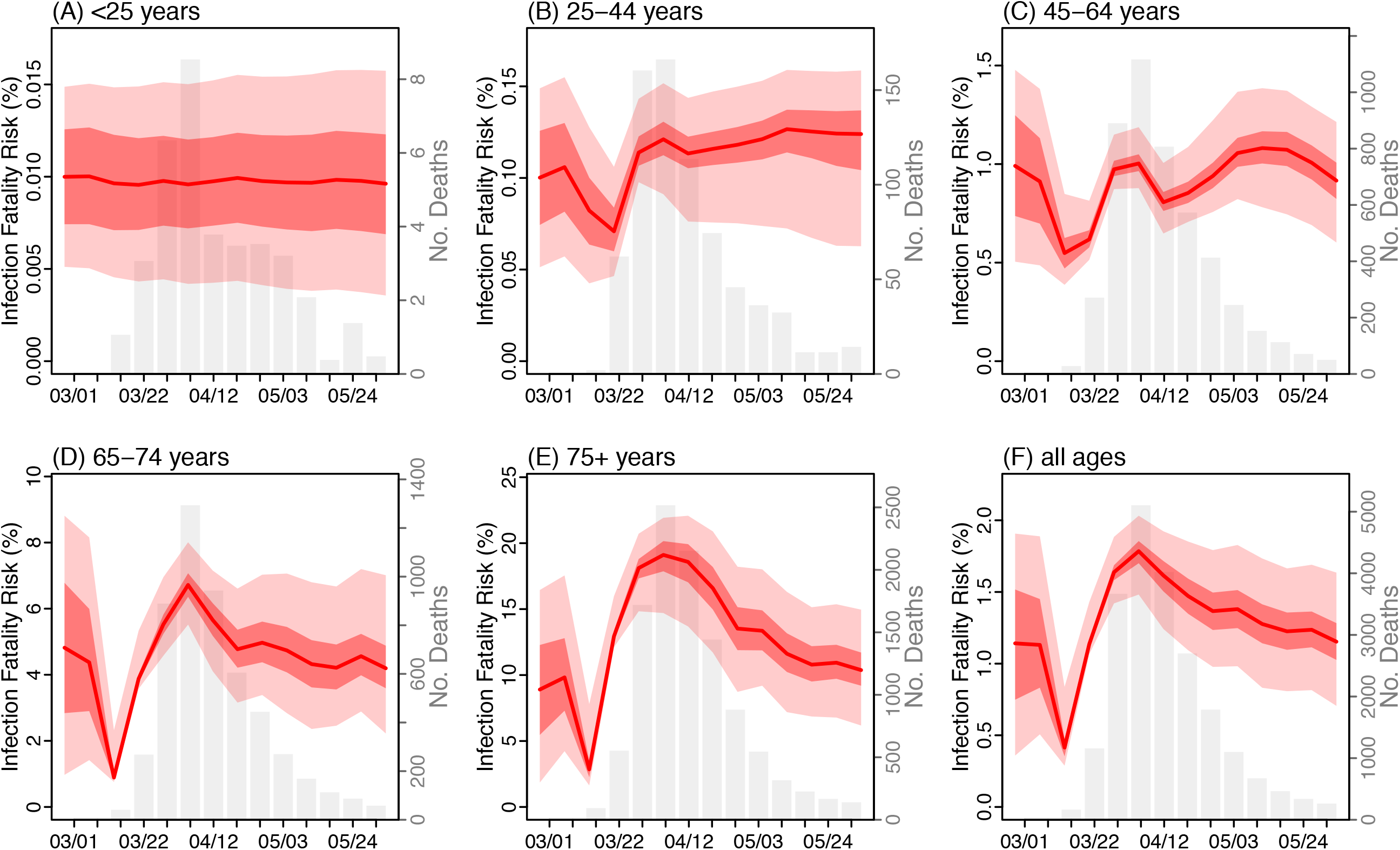
Estimated infection fatality risk for (A) <25 year-olds, (B) 25-44 year-olds, (C) 45-64 year-olds, (D) 65-74 year-olds, (E) 75+ year-olds, and (F) all ages overall. Red lines show the estimated median IFR with surrounding areas indicating the 50% and 95% CrI. For comparison, the grey bars show the number of deaths reported for each week from the week of March 1 to the week of May 31, 2020.

## Discussion

In light of the large uncertainties in IFRs for COVID-19 due to under-detection of infections, we have used a model-inference system, developed to support the pandemic response in NYC, to estimate local IFRs. During the 2020 spring pandemic (March 1–June 6, 2020), NYC recorded the largest numbers of COVID-19 cases and deaths in the US and perhaps worldwide. Despite public health efforts to slow the pandemic, e.g. by social distancing, and to ramp up healthcare capacity, over 21,000 lives were lost from COVID-19 in a short span of three months. Based on this large number of deaths, the estimated overall IFR was 1.39% in NYC. This estimate included both confirmed and probable COVID-19 deaths. If only COVID-19 confirmed deaths were included, given that 78.9% of deaths among the total were laboratory-confirmed, the estimated overall IFR would be around 1.1%. Both estimates were higher than previously reported for elsewhere (e.g., about 0.7% in both China^3^ and France^5^). Importantly, NYC has nosologists who review all death certificates and record deaths into a unified electronic reporting system rapidly (the average death certification time was 22.2 hours and 95% of deaths were certified within 3.1 days during the pandemic wave). This mortality surveillance infrastructure and enhanced nosology thus allow more rapid and complete death reporting in NYC. As such, our estimates here likely more accurately reflect the underlying fatality risk of COVID-19 infection. Further, given the likely stronger public health infrastructure and healthcare systems in NYC than many other places,^18^ the higher IFR estimated here suggests that fatality risk from COVID-19 may be higher in the United States and likely other countries as well than previously reported. Of note, despite the large surge in cases and hospitalizations, through quick expansion of healthcare systems, most hospitals in NYC were able to meet patient care demand during the pandemic.

As COVID-19 continues to pose pandemic risk in many places worldwide, it is crucial that governments account for and closely monitor the infection rate and health outcomes including hospitalizations and mortality and take prompt public health responses accordingly.

While the IFR estimated here was similar to those previously reported elsewhere for younger age groups, we found that IFRs for individuals 65 years and older in NYC were about twice as high as prior reports.^3^ These higher IFRs may be in part due to differences in population characteristics, in particular, the prevalence of underlying medical conditions such as diabetes mellitus, chronic lung disease, and cardiovascular disease.^19,20^ Regardless, estimated weekly IFR was as high as 6.7% for 65-74 year-olds and 19% for 75+ year-olds. These dire estimates highlight the severity of COVID-19 in elderly populations and the importance of infection prevention in congregate settings. Thus, early detection and adherence to infection control guidance in long-term care and adult care facilities should be a priority for COVID-19 response as the pandemic continues to unfold.

In addition, over 5000 COVID-19 deaths also occurred among younger adults aged 25-64 years during the pandemic (734 deaths among 25-44 year-olds and 4732 deaths among 45-64 year-olds; Table 1). Despite this large number of COVID-19 deaths, estimated cumulative infection rates in these age groups were only around 20% by the week of May 31, 2020, much lower than the 50-70% herd immunity needed to prevent large epidemics of COVID-19 (assuming the basic reproductive number for SARS-CoV-2 is around 2-3.5 and infection confers long-lasting immunity).^5,21,22^ By July 2020, many places where lockdown-like measures were lifted saw increases in COVID-19 cases among young adults.^23-25^ These continuous infections could ignite new epidemics of COVID-19 and lead to further devastating impacts in elderly populations as well as younger adults (in particular, those aged 45-64 years), given the remaining high population susceptibility in many places and transmission across age groups. As such, it is essential that young adults strictly adhere to social distancing and preventive measures (e.g., mask wearing) in places with continuous transmission, despite their relatively low IFRs.

In this study, we incorporated multiple data sources, including age-grouped, spatially resolved case and mortality data as well as mobility data, to calibrate our model-inference system. Of note, the timing of the COVID-19 pandemic varied substantially among NYC neighborhoods. For instance, peak mortality rates occurred up to 4 weeks apart among the 42 neighborhoods. Fitting the model-inference system simultaneously to these diverse case and mortality time series thus enabled better constraint of key model parameters (e.g., infection detection rate and IFR). However, we note there remain large uncertainties in model estimates. A full assessment of COVID-19 severity will require comprehensive serology surveys of the population by age group and neighborhood, given the large heterogeneity of infection rates across population segments and space. In addition, we only included deaths that were lab-confirmed or explicitly coded as related to COVID-19. Previous studies have reported that excess deaths in NYC during about the same period were more than 24,000.^8,26^ Further, recent studies have reported severe sequelae of COVID-19 in children, i.e. Multi-system Inflammatory Syndrome in Children. Thus, it is important to monitor health outcomes in younger age groups post-infection as the pandemic unfolds, despite the low IFRs noted to date.

## Data Availability

COVID-19 case and mortality data for New York City publicly available from https://github.com/nychealth/coronavirus-data

## Acknowledgments

This study was supported by the National Institute of Allergy and Infectious Diseases (AI145883), the National Science Foundation Rapid Response Research Program (RAPID; 2027369), and the NYC DOHMH. We thank the NYC DOHMH Bureau of Vital Statistics team and staff members in the NYC DOHMH Incident Command System Surveillance and Epidemiology Section for data management. We thank Miranda S. Moore at NYC DOHMH, who wrote and maintained the code to generate weekly extracts of COVID-19 case data for provision to Columbia University under a data use agreement. We thank Jaimie Shaff at NYC DOHMH and Helen Alesbury and Kathryn Klein at the Office of Chief Medical Examiner of the City of New York for initiating and coordinating discussions on modeling COVID-19 mortality in NYC. We also thank Columbia University Mailman School of Public Health for high performance computing and SafeGraph (safegraph.com) for providing the mobility data used in this study.

## Appendix

This document includes: 1) A comparison of model estimated cumulative infection rates with measured seroprevalence of antibodies to SARS-CoV-2, for independent model validation; 2) Supplemental method and results for estimating the potential seasonality of SARS-CoV-2 in New York City; and 3) Supplemental tables and figures.

### 1. Comparison of model estimated cumulative infection rates with measured seroprevalence

There are large uncertainties in our model estimates, in particular, the estimated cumulative infection rates. As a model validation, we compare our estimates of cumulative infection rates to available data based on measured seroprevalence of antibodies to SARS-CoV-2. We identified three sets of seroprevalence data for three key time points of the pandemic period:

1. Reports from Havers et al. 2020,^1^ by mid-March (i.e., the early phase of the pandemic wave);
2. Data release from the New York State Governor’s COVID-19 daily briefing,^2^ by mid-April (i.e., around the peak of the pandemic wave); and
3. Data release from the New York State Governor’s COVID-19 daily briefing,^3^ by early June (i.e., end of the pandemic wave)

Table S2 summarizes the serology data, including the timing of measurement, sampled population and potential biases, age groups, and geographic locations. For the comparison, we matched our model estimates by timing of measurement (assuming a lag of ~10 days between infection and the development of antibodies), age groups, and geographic locations (either citywide or by borough).

Figure S1 overlays the model estimated cumulative infection rates and corresponding seroprevalence measures, stratified by age group and location. Taking into account potential biases in the serology data (i.e., likely higher than the general population due to sampling methods; Table S2), our model estimates were in line with the seroprevalence measures for different age groups as well as for different geographic locations, at all three time points.

### 2. Estimating the potential seasonality of SARS-CoV-2 in New York City

Respiratory viruses such as influenza and human endemic coronaviruses tend to cause infections in humans predominantly in cold months in temperate regions. Similarly, SARS-CoV-2 transmission could share such a seasonal pattern. Therefore, here we used routine viral surveillance data for human coronaviruses collected in New York City during 2015-2019 to estimate the potential seasonality for SARS-CoV-2. There are four endemic coronaviruses infecting humans (OC43, 229E, NL63, HKU1). Of these four viruses, OC43 and HKU1 are betacoronaviruses, i.e. in the same genus as SARS-CoV-2; in addition, OC43 circulates most frequently in humans in NYC (Fig S2). Therefore, we estimated the seasonal cycle based on two coronavirus time series: 1) combining data for all four human endemic coronaviruses; and 2) OC43 alone.

#### 2.1 Data processing

The viral surveillance data included the number of respiratory samples collected and the number tested positive for each coronavirus, respectively, during each week from the Week of Oct 4, 2015 to the Week of Feb 9, 2020 (i.e. before the COVID-19 pandemic). To adjust for fluctuations in number of samples collected over time, we scaled the number of positive samples for each week by a factor of *p*, where *p* = total number of samples collected during the 2015-2016 season (i.e., the first season of the dataset) / total number of samples collected during the corresponding season. A season was defined as Morbidity and Mortality Weekly Report (MMWR) Week 40 of this year to MMWR Week 20 of the following year (i.e., the same as for the influenza season per the United States Centers for Disease Control and Prevention). For the combined coronavirus time series, we computed the number of samples tested positive (after the scaling) for any human endemic coronaviruses for each week. To smooth the weekly data, we further computed the 3-week centered moving average for each time series and used the smoothed time series for estimating the seasonal cycle.

#### 2.3 Modeling the seasonal cycle

To estimate the seasonal cycle, we built a model-inference system using a susceptible-exposedinfectious-recovered-susceptible (SEIRS) model in conjunction with a particle filter with space reprobing.^4,5^ The SIRS model took the following form:

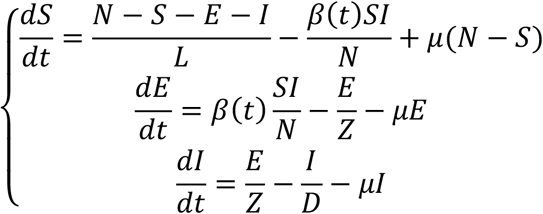

where *S, E*, and *I* are the number of susceptible, exposed, and infectious individuals, respectively; *N* is population size; therefore, *N* – *S* – *E* – *I* is the number of recovered individuals. *μ* is the birth rate and death rate (i.e. set to the same value to keep population size constant; here *μ* was set to 13.6/1000 per year for NYC). *β*(*t*) is transmission rate during week-*t*, which can vary over time due to seasonality. *Z, D* and *L* are the latency, infectious, and immunity period, respectively. The basic reproductive number *R*_0_(*t*) is computed as *R*_0_(*t*) *= β*(*t*)*D*.

To estimate model parameters, *β*(*t*)*, D* and *L*, for each week, we ran the SEIRS model *stochastically* in conjunction with a particle filter with space reprobing and fit to the weekly coronavirus time series (either for all coronaviruses combined or OC43 alone; scaled by a factor of 10 to account for under-detection) from the Week of Oct 4, 2015 to the Week of Feb 9, 2020. The particle filter uses a suite of model realizations (*n*=5000 here), each with initial parameters and variables randomly drawn from a prior range. After model initiation, the model is integrated one time step (i.e. 1 week here) forward, which generates the prior for that time step; the filter then computes the posterior for that time step per Bayes’ rule and resamples the parameters to update the parameter distribution. This filtering cycle is repeated until the last data point is incorporated.

We draw the initial parameters using the following uniform distributions: U[0.1, 0.8] per day for *β*, U[2, 3] days for *Z*, U[4, 6] days for *D*. For the immunity period *L*, due to the large uncertainty, we tested multiple combinations of *L* and susceptibility *S*. For all coronaviruses combined, we found that the system was most stable when *S* was 50-60% of the population and *L* was around ~3 years (see Figure S3); therefore, we used the following initial ranges: U[50%, 60%] of the population for *S* and U[900, 1100] days for L. For OC43, we found that the system was most stable when *S* was 60-70% of the population and *L* was around ~5 years (see Figure S4); therefore, we used the following initial ranges: U[60%, 70%] of the population for *S* and U[1800, 2200] days for L. To account for stochasticity in model initiation, we ran the parameter estimation process independently 5 times.

Using the posterior estimates of *R_0_* for each week, we computed the weekly average for each week of the year (e.g., *R_0_* for Week 40 was the average of all available estimates for the same week) and further smoothed the weekly averages using 3-week centered moving average. Finally, we scaled the smoothed weekly averages by their mean to generate the relative *R_0_* seasonal cycle. The relative *R_0_* seasonal cycle was used to adjust the weekly *R_0_* estimates for SARS-CoV-2. For instance, estimated relative *R_0_* during the first week of the year was 1.2; the *prior* of the transmission rate (and thus *R_0_*) for the same week was multiplied by 1.2 to account for the more conducive transmission conditions at that time.

#### 2.4 Results

Figure S1 shows the seasonal pattern for the four human endemic coronaviruses individually and combined. Figure S3 (all human coronaviruses combined) and Figure S4 (OC43) show the model-fit and posterior estimates of population susceptibility and *R_0_* during Oct 4, 2015 – Feb 9, 2020. Figure S5 shows the estimated *R_0_* seasonal cycle based on the two human coronavirus time series, separately. The estimated *R_0_* seasonal cycles were similar for the two time series. We thus used the one based on OC43 (in the same genus as SARS-CoV-2) to represent the seasonality of SARS-CoV-2 for the full model-inference system reported in the main text. We note, however, that these results remain preliminary.

## Supplementary Tables

**Table S1.**
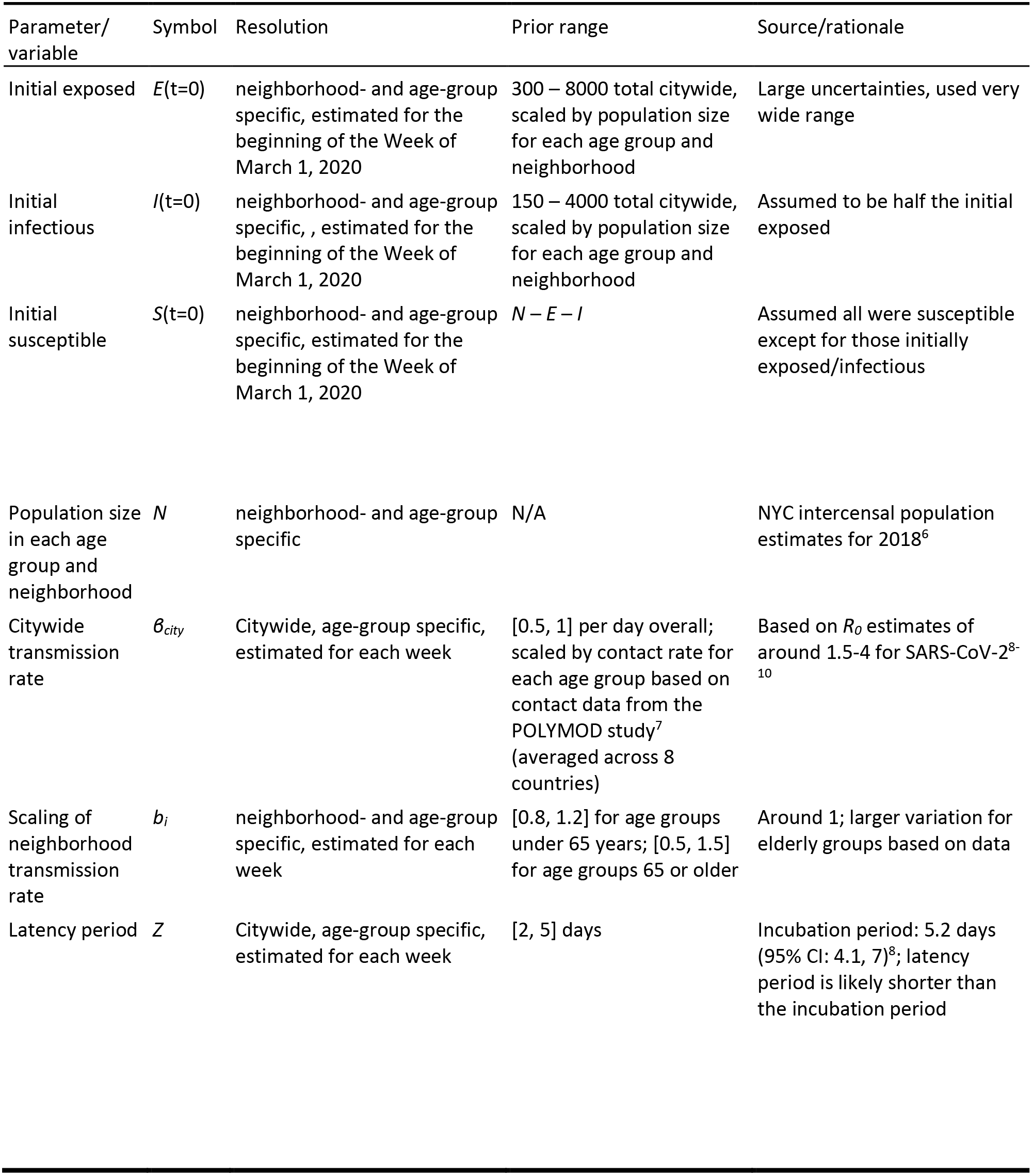
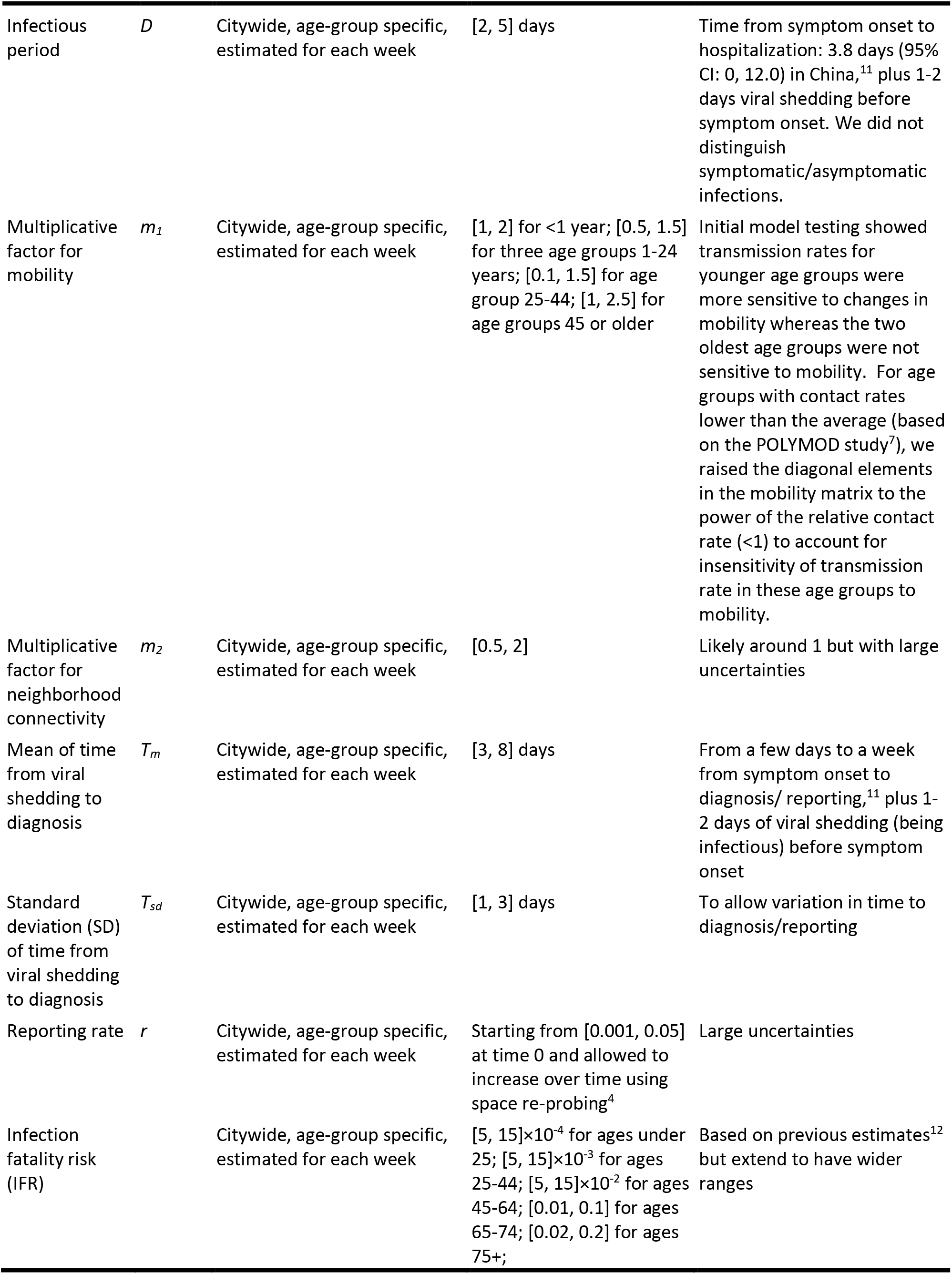
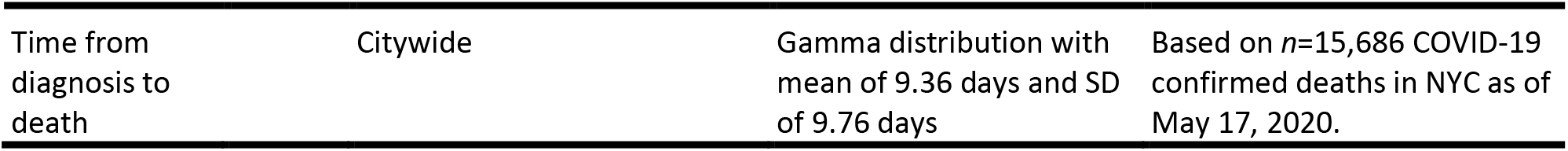
Prior ranges for main model parameters and variables. The spatial, temporal, and age resolution of each parameter or variable, estimated in the model-inference system, is specified in the column “Resolution”. Note posterior parameter estimates can extend outside the specified prior ranges.

**Table S2.** Summary of serology studies and data used for comparison with model estimates.

| Serology surveys | Matching Study period |  | Sample/survey population | Location of sample collection/survey | Potential biases of serology data | Matching age group |  |
| --- | --- | --- | --- | --- | --- | --- | --- |
|  | serology data collection | model estimates |  |  |  | serology data | model estimates |
| Havers et al. 2020<sup>1</sup> | March 23 – April 1, 2020 (sample collection dates) | Week ending March 21, 2020 | Convenience samples from clinics<sup>1</sup> | Mainly Manhattan, Bronx, Brooklyn, Queens, and Nassau counties | Potentially from patients seeking COVID-19 treatment<sup>1</sup> and thus higher positivity rates than the general population | 0-18 years | <25 years |
|  |  |  |  |  |  | 19-49 years | 25-44 years |
|  |  |  |  |  |  | 50-64 years | 45-64 years |
|  |  |  |  |  |  | >65 years | 65-74, 75+ years |
|  |  |  |  |  |  | all | all |
| Governor 's COVID-19 daily briefing<sup>2</sup> | May 1, 2020 (report date) | Week ending April 18, 2020 | NYC residents visiting grocery and big-box stores.<sup>13</sup> | Citywide and for individual boroughs: Manhattan, Bronx, Brooklyn, Queens, Staten Island | Potentially from adults 25-64 year-olds who were outside during the pandemic and thus higher positivity rates than the general population | Not reported; likely 25-64 years | 25-44 years<br>45-64 years<br>all |
| Governor 's COVID-19 daily briefing<sup>3</sup> | June 13, 2020 (report date) | Week ending June 6, 2020 (i.e. end of our study period) | Likely NYC residents visiting grocery and big-box stores.<sup>13</sup> | Citywide and for individual boroughs: Manhattan, Bronx, Brooklyn, Queens, Staten Island | Potentially from adults 25-64 year-olds who were outside during the pandemic and thus higher positivity rates than the general population | Not reported; likely 25-64 years | 25-44 years<br>45-64 years<br>all |

## Supplementary Figures

**Figure S1.**
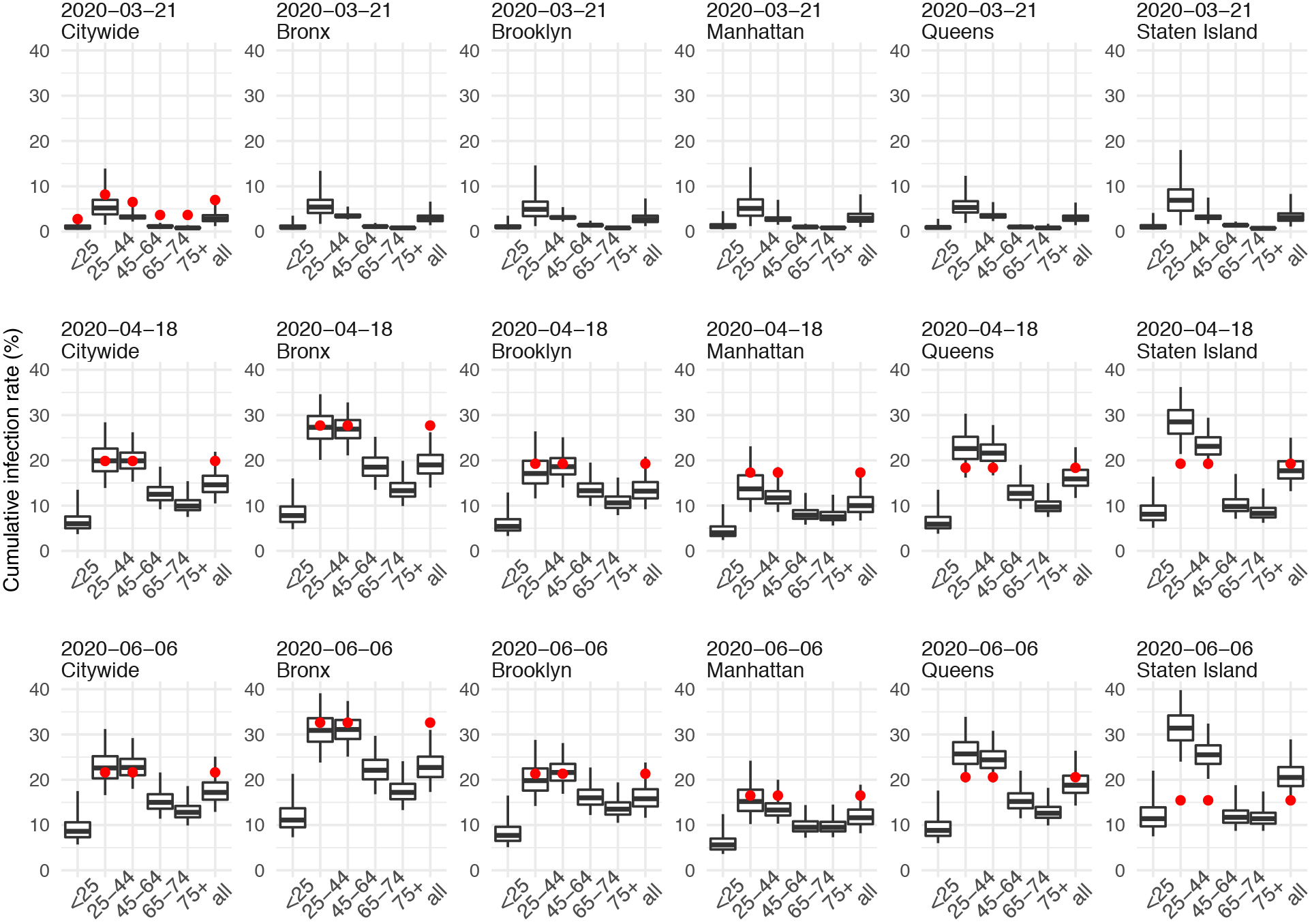
Model validation. We compare model estimated cumulative infection rates with available seroprevalence measures at three time points of the pandemic wave: the week ending March 21 (early phase of the pandemic; top panel), the week ending April 18 (around the peak; middle panel), and the week ending June 6 (end of the pandemic; bottom panel). Boxes show model estimated cumulative infection rates up to the specified dates, stratified by age group and location. Thick horizontal lines and box edges show the median, 25^th^, and 75^th^ percentiles of model estimates; vertical lines extending from each box show 95% Crl. Red dots show measured seroprevalence of antibodies to SARS-CoV-2 during the corresponding time period (see details on the serology data and matching of study period and age group in Table S2).

**Figure S2.**
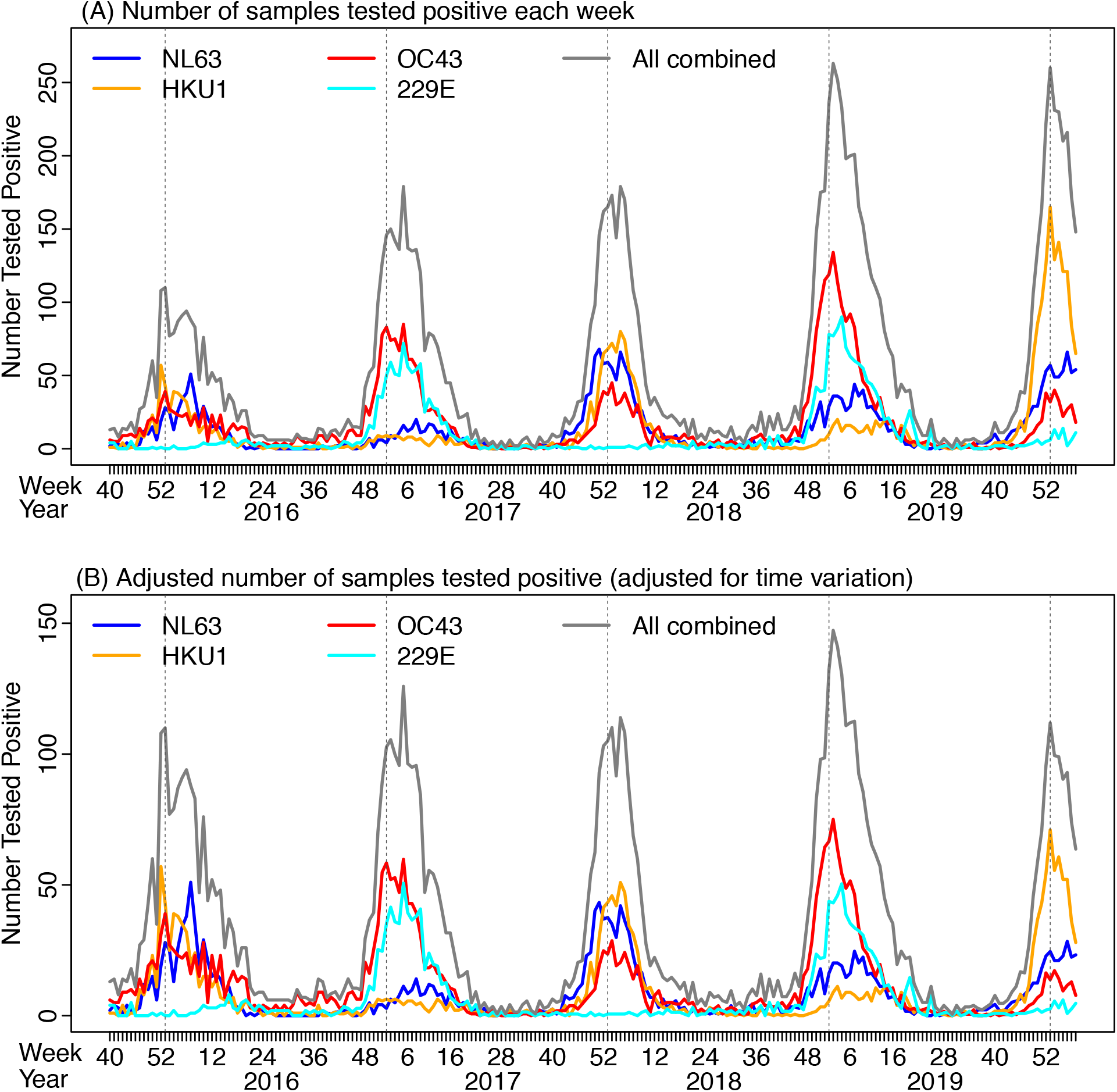
Human endemic coronavirus viral surveillance data collected in NYC. Lines show the number of respiratory samples tested positive for different coronaviruses without adjustment (A) or after adjusting for increasing sample collection (B) during Oct 4, 2015 – Feb 9, 2020.

**Figure S3.**
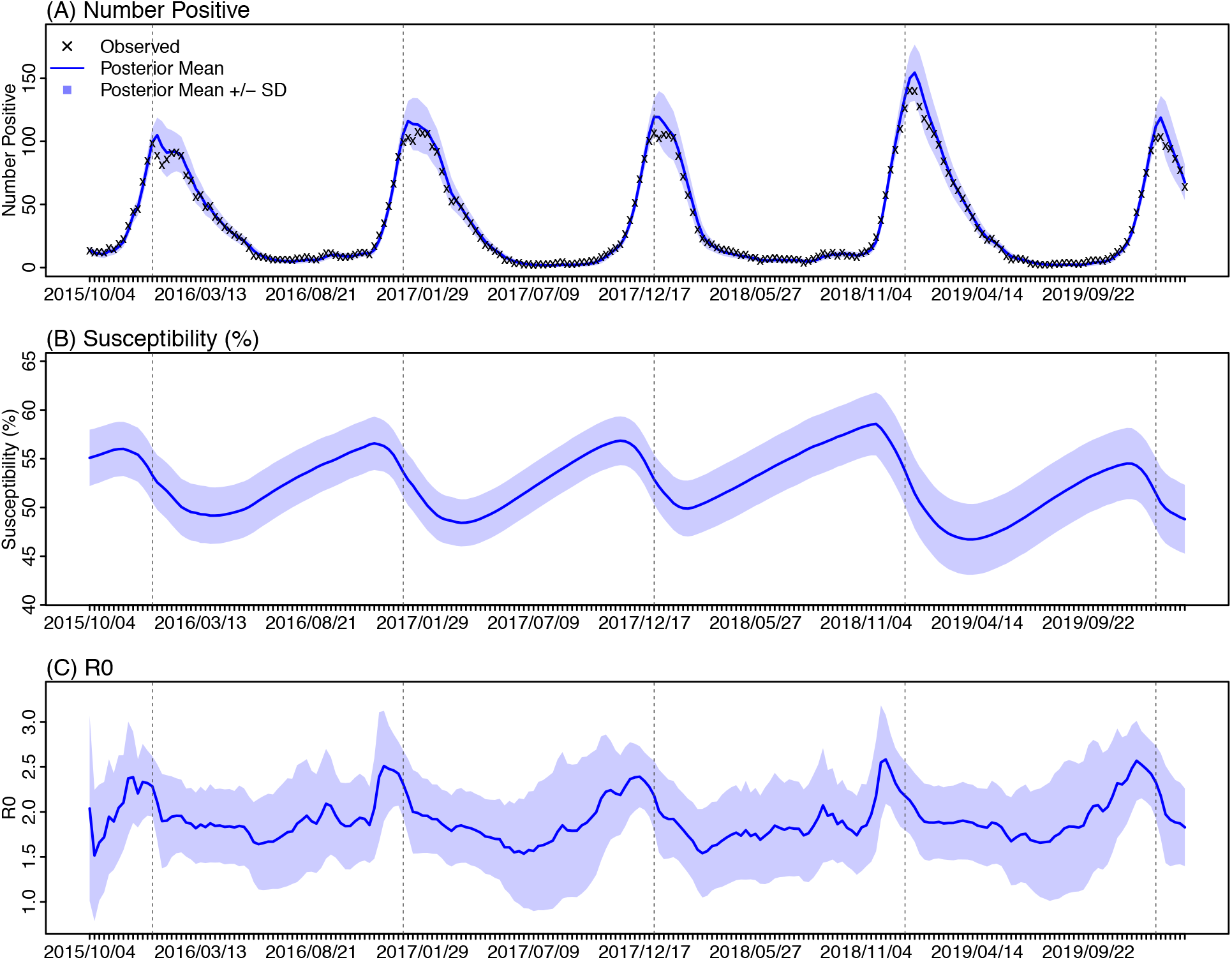
Model-fit and key parameter estimates for all human coronaviruses combined: (A) Model fit to the time series; (B) Weekly posterior estimates of corresponding population susceptibility; and (C) Weekly posterior estimates of *R*_0_. ‘x’s show observations; blue lines show posterior mean estimates and shaded areas show posterior mean ± 1 standard deviation (SD).

**Figure S4.**
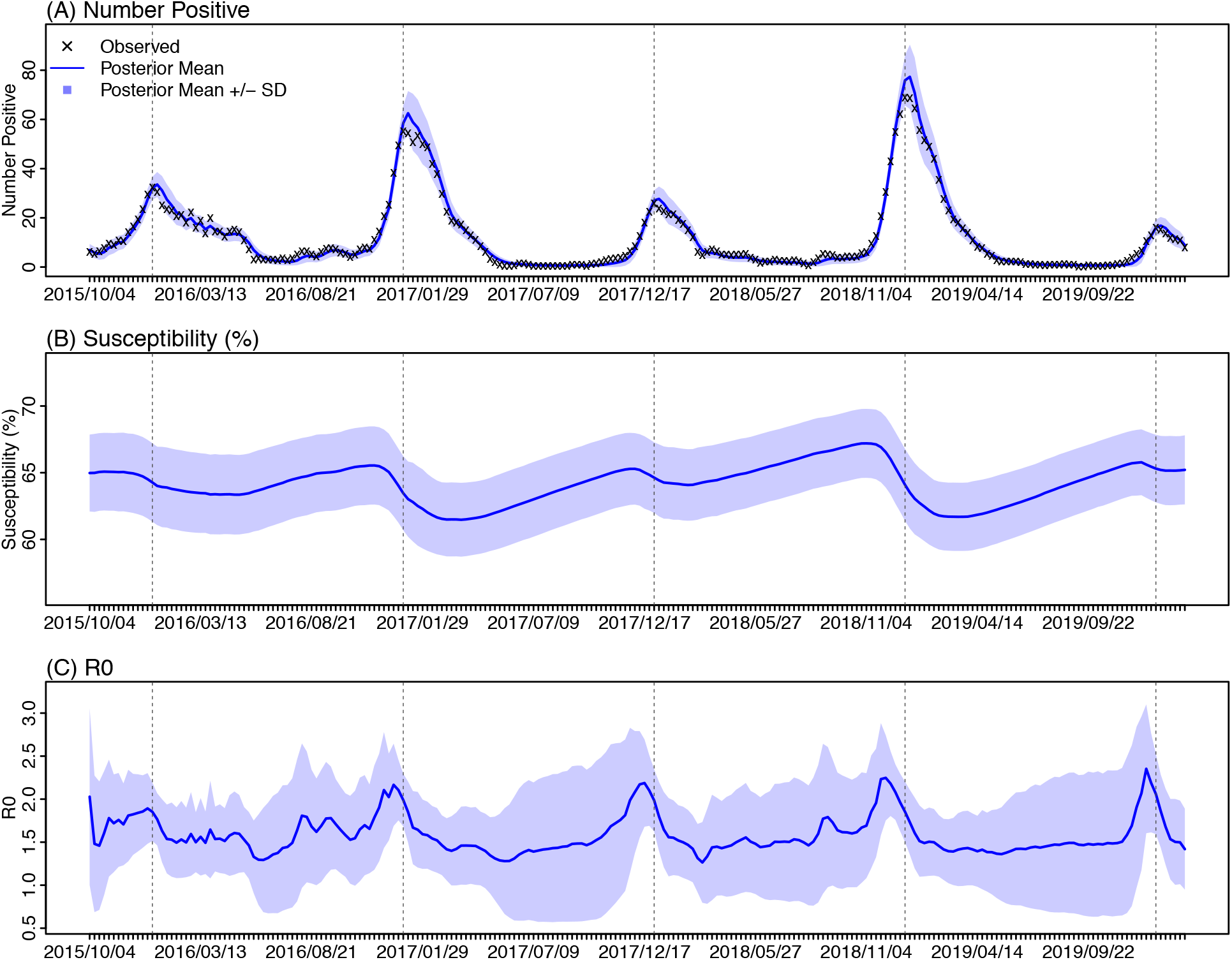
Model-fit and key parameter estimates for OC43: (A) Model fit to the time series; (B) Weekly posterior estimates of corresponding population susceptibility; and (C) Weekly posterior estimates of *R*_0_. ‘x’s show observations; blue lines show posterior mean estimates and shaded areas show posterior mean ± 1 standard deviation (SD).

**Figure S5.**
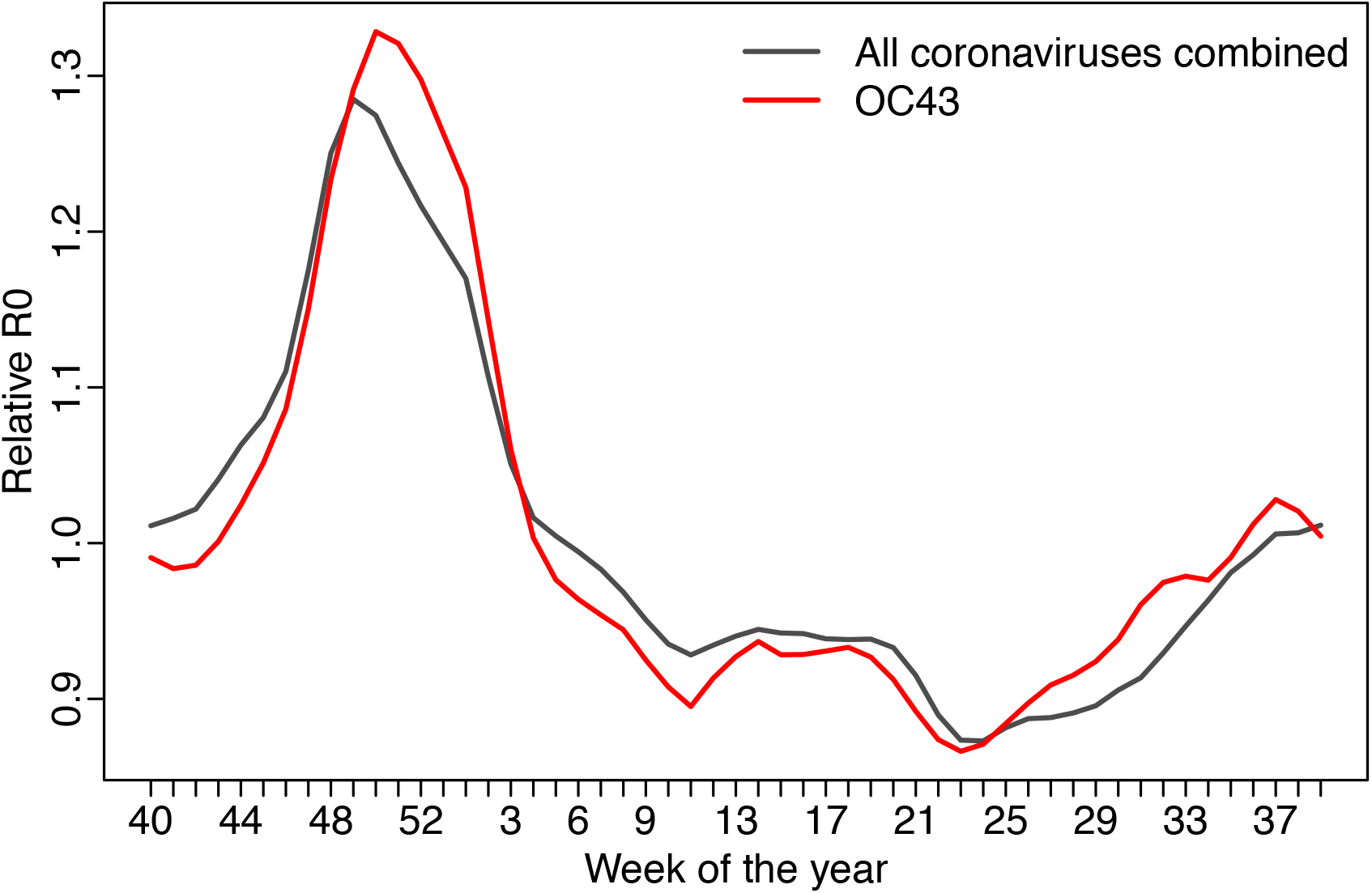
Estimated seasonal cycle. Lines show the R_0_ estimate relative to the yearly mean for each week of the year.

